# A Phase I trial to inform clinical protocols for the safe administration of psilocybin-assisted psychotherapy

**DOI:** 10.1101/2023.04.12.23288325

**Authors:** Jennifer N. Bennett, Michael D. Blough, Ian Mitchell, Lyle Galloway, Ravinder Bains

**Author notes:** These authors contributed equally and share first authorship (Jennifer N. Bennett, Michael D. Blough, Ravinder Bains). **Correspondence:** Jennifer Bennett.

## Abstract

This Phase I trial aims to inform the development of safety protocols for psilocybin-assisted therapy. Psychedelics, including psilocybin, are increasingly being recognized as a successful treatment option for many mental health concerns. In order to decrease the risks associated with its clinical use, more data is required regarding its physiological effects in healthy individuals. Safety assessments (heart rate, blood pressure, temperature, and ECG data), as well as adverse event evaluations were the primary outcome measures used to assess the physiological effects of 25 mg of psilocybin extract administered to 14 healthy individuals. We hypothesized that there would be a transient, clinically insignificant rise in both blood pressure and heart rate that would not result in any long-term adverse effects. No unexpected effects were observed, blood pressure and heart rate returned to normal as drug effects waned, and all participants had normal two-month follow-ups. Mean peak systolic and diastolic blood pressures during the psilocybin session were 145.93 (*SD* = 19.01) and 93.93 (*SD* = 9.75), respectively. While this represents a significant increase from baseline (*p* < 0.0001), a healthy cardiovascular system is capable of tolerating such levels for a longer time period than the brief duration of drug effects. Therefore, we suggest implementing focused and limited screening protocols to balance patient safety and accessibility. Secondary outcomes of this trial centered on the subjective effects of psilocybin, assessed via the QIDS-SR16 and the MEQ-30. There was a statistically significant decrease in QIDS-SR16 scores from baseline scores (*M* = 3.50, *SD* = 2.35) to eight-week follow-up scores (*M* = 1.86, *SD* = 0.86), *p* = 0.018. Mean MEQ-30 scores, assessed on day two and seven after the psilocybin session, indicate participants had full mystical experiences.

## Introduction

Over the last two decades, research on psychedelic drugs for the treatment of mental health disorders has increased exponentially. Psilocybin – the psychoactive compound in hallucinogenic mushrooms that are largely of the genus *Psilocybe* – has been gaining prominence in the treatment of generalized anxiety disorder, treatment-resistant depression, and end-of-life anxiety in palliative care patients (Carhart-Harris & Goodwin, 2017; Siegel et al., 2021). Worldwide, countless studies have been conducted examining the efficacy of psilocybin in treating such disorders, with many more clinical trials currently underway to assess its efficacy in managing other indications such as eating disorders, post-traumatic stress disorder (PTSD), substance use disorders, obsessive-compulsive disorder (OCD), chronic pain, and migraines. While its efficacy has been broadly accepted by now, further exploration is nonetheless warranted for potential contraindications and adverse events that psilocybin may induce.

Psilocybin is classified as a Schedule I drug in the United States (DEA, 2020); this classification has largely influenced the stigma that it has high abuse potential, no accepted medical use, and can result in serious adverse events. However, research demonstrates contrary findings, insofar as psilocybin not only has a low abuse potential but can actually aid in substance abuse cessation and treat multiple mental health disorders, while maintaining a relatively positive safety profile (Lowe et al., 2021). Nonetheless, as with any medication, establishing proper screening and safety protocols is critical in order to implement this therapy responsibly.

When used properly, psilocybin has a favourable risk-benefit ratio. Mental illnesses are devastating disorders that carry the potential risk of harm to self and harm to others, both directly and indirectly; psilocybin, on the contrary, has a high therapeutic index of 641 (a better safety profile than both aspirin and nicotine), and has significant potential to decrease the burden of these disorders (Lowe et al., 2021). Although generally considered physiologically safe and psychologically beneficial, it is important that clients are supervised during the acute psychedelic experience so that support can be offered should any potential anxiety, trauma, impaired decision-making, and/or rare physiological side effects arise. As with any pharmaceutical, there are certain concomitant medications and medical conditions that should be avoided in order to warrant the drug’s safe use. There is the need to gather more data examining baseline physiology in healthy individuals to demonstrate that psilocybin can be safely administered in a clinical setting. From the results obtained in our Phase I safety trial, we aim to inform the development of protocols for psilocybin-assisted psychotherapy.

Vital signs and adverse event assessments were the primary outcome measures in this trial. Such assessments can help identify which populations present with an elevated risk, and thus for whom extra caution would be advisable. A proper screening protocol can then be developed from the evaluation of adverse events. Several studies have previously assessed the cardiovascular effects of psilocybin, in which transient increases in blood pressure were observed (Hasler et al., 2004; Daniel & Haberman, 2017; Carbonaro et al., 2018). Hasler et al. (2004) also showed that ECG was unaffected with various doses of psilocybin; however, Dahmane et al. (2021) showed a minor effect of 25mg on QTc prolongation, in which the effect of 25mg of psilocybin meets a low level of concern (<10 ms) regarding non-cardiac medications by the FDA. As such, this trial sought to further evaluate sympathomimetic effects including heart rate, blood pressure, cardiac electrical activity via ECG, and temperature. Adverse event assessments also included monitoring for side effects for up to two months after the psilocybin session.

The secondary exploratory objective of this study examined the effects of psilocybin on both mood and mystical experience. Two questionnaires – the Quick Inventory of Depressive Symptomatology Self-Report 16-item (QIDS-SR16) and the 30-item Revised Mystical Experience Questionnaire – were used as the assessment tools to measure these variables.

## Materials and Methods

### Study Design

This was an open label, single arm, single centre Phase I safety trial that took place at ATMA Journey Centers in Calgary, Canada. Participants were recruited nationally, and during the enrollment process, participants were screened by the Primary Investigator for eligibility based on the criteria approved by Health Canada. Participants were followed for two months after the psilocybin session. This trial was approved by The Health Research Ethics Board of Alberta (HREBA) – Clinical Trials Committee (CTC).

### Investigational Product

The psilocybin product, PEX010, was provided by Psilo Scientific Ltd. in the form of psilocybin standardized extract powder, encapsulated in hydroxypropyl methylcellulose capsules. Each capsule contained 25 mg of psilocybin and was manufactured in full compliance with Good Manufacturing Practices (GMP) required to manufacture investigational medicinal products in Canada.

### Participants

During screening, applicants were evaluated for eligibility criteria, in which 14 participants were selected. The criteria required that participants be aged 18-65 years of age, be physically and mentally healthy as determined by a primary care physician via physical and mental examinations, be a medical or mental healthcare provider with professional accreditation, have a negative pregnancy test at study entry and prior to the psilocybin session if of child-bearing potential, and agree to use adequate forms of birth control from study entry to 10 days after the psilocybin session. Exclusion criteria included both active psychotic symptoms and a history of psychotic symptoms, bipolar disorder, schizophrenia, first-or second-degree relatives with a history of psychotic symptoms, bipolar disorder, or schizophrenia, the use of psychotropic medications including SSRIs, SNRIs, MAOIs, or lithium, a diagnosis of either dementia or delirium, a high risk for coronary artery disease, uncontrolled cardiopulmonary disease/cardiovascular disease, uncontrolled hypertension greater than 140/90mmHg assessed on three separate occasions, a history of QT prolongation or on concomitant medications carrying a risk of QT prolongation, aneurysm, a history of intracerebral hemorrhage, hepatic cirrhosis, hepatorenal disease, any other clinically significant medical condition or disease, suicide risk, or have a known sensitivity or intolerability to psilocybin or its metabolites. This eligibility criteria were developed based on previously researched contraindications and other potential contraindications that require further research before psilocybin can comfortably be used by such populations. Participants were required to sign an informed consent form before enrollment.

The criterion that participants must hold professional accreditation as a medical or mental healthcare provider produced a unique opportunity to examine the effectiveness of psilocybin in a clinical setting; healthcare workers who work directly with patients suffering from mental illness are likely to be valuable judges of a treatment’s utility and possess a constructive perspective. Although not a measurable objective in this study, it gives the professionals involved in the development of protocols and the implementation of this novel treatment a practical understanding of how and why it is efficacious, as well as laying the foundation for future research examining this as a formal objective.

As shown in Table 1, this study enrolled 14 participants aged 32-64. Participants were predominately female (78.6%), with 77% of total applicants being female. Although this is disproportionate, an analysis performed by the World Health Organization including 104 countries showed that 70% of the total population of workers in the health and social sector are female (WHO, 2019).

**Table 1:** Participants’ demographic information.

| Characteristic | n = 14 |
| --- | --- |
| **Age**, range (mean, SD) | 32-64 (46, 10.33) |
| **Gender**, n (%) |  |
| Female | 11 (78.6) |
| Male | 3 (21.4) |
| **Designations**, n (%) |  |
| Registered Nurse | 3 (21.4) |
| Registered Psychiatric Nurse | 1 (7.1) |
| Occupational Therapist | 1 (7.1) |
| Registered Social Worker | 3 (21.4) |
| Registered Psychologist | 4 (28.6) |
| Registered Clinical Counsellor | 1 (7.1) |
| Registered Psychotherapist | 1 (7.1) |

### Outcome Measures

The primary objective of this Phase I clinical trial was to collect data regarding the safety of psilocybin when administered to healthy individuals. Safety was assessed by documenting vital signs and recording adverse events (AEs) and serious adverse events (SAEs).

The secondary exploratory objective utilized two self-assessment questionnaires (the QIDS-SR16 and the MEQ-30). These questionnaires were intended to evaluate the general nature of the subjective effects occasioned by the consumption of psilocybin and to determine its impacts on the participants’ mental health following their experimental session.

### Safety Assessments

For safety evaluations, SAEs (defined as any untoward medical occurrence that happens after psilocybin treatment that either results in death, is life-threatening, requires in-patient hospitalization, results in persistent or significant disability/incapacity, results in a congenital anomaly or birth defect, or a loss of pregnancy) and all adverse events (defined as unfavourable and unintended signs, symptoms, or disease associated with a treatment) were formally assessed during the psilocybin session, as well as two days, seven days, and eight weeks following the psilocybin session. During this eight-week time period, participants were also instructed to report any SAEs immediately. AEs were assessed by severity (mild, moderate, severe, life-threatening, or fatal). Specific AEs that were investigated include nausea, vomiting, headache, anxiety, confusion, fatigue, and mania or psychotic symptoms. A subset of AEs – adverse events of special interest (AESI) –that could be indicative of QT interval prolongation or cardiac arrhythmias (including ECG abnormalities during the psilocybin session, sudden death, non-postural syncope, palpitations, or seizures) were assessed to evaluate psilocybin’s impact on cardiac function. A second AESI involved the assessment of suicide risk via the following behaviours: suicide, suicide attempts, self-injurious behaviour associated with suicidal ideation, or suicidal ideation judged to be serious or severe in the opinion of the Principal Investigator.

Treatment emergent adverse events (TEAEs; defined as AEs with an onset after drug administration and up to two days afterwards) were also assessed by severity, and included assessments for nausea, vomiting, headache, anxiety, confusion, fatigue, and mania or psychotic symptoms that emerged soon after the psilocybin session and lasted for longer than 24 hours after cessation of drug effects. Adverse drug reactions (ADRs; defined as a serious adverse event that is not identified in nature, severity, or frequency in the risk information set out in the product monograph) were monitored, with special attention being paid to unexpected ADRs.

During the psilocybin session, blood pressure, heart rate, and temperature were measured every hour, as well as when signs or symptoms warranted further vital sign measurement. 30-second electrocardiogram (ECG) recordings were obtained every hour. Blood pressure and heart rate were monitored using the Large Cuff Easy@Home Digital Upper Arm Blood Pressure Monitor, FDA-cleared for over-the-counter use. Temperature was assessed via the FOR A IR42 Forehead Thermometer, licensed by Health Canada. ECG recordings were obtained using the KardiaMobile Six-Lead Personal EKG Monitor.

### Statistical Analysis

The statistical analysis was completed using SPSS 29.0 and Prism 9.

## Results

Table 2 summarizes the main results of this study. A t-test for dependent means was used to assess the effect that psilocybin had on peak mean arterial pressure (MAP). To do this, a corresponding MAP was calculated from each blood pressure measurement that was taken during the psilocybin session, using the formula MAP = DP + 1/3(SP – DP). The highest MAP of each participant was documented and used to help determine the risk psilocybin poses for the cardiovascular system. Figure 1 compares baseline to peak MAP. There was a statistically significant effect *t*(13) = -5.56, *p* < 0.001, of psilocybin on MAP. Mean arterial pressure increased from an average baseline of 91.55 mmHg (*SD* = 7.59) to an average peak of 110.76 mmHg (*SD* = 12.15) during the psilocybin session. Baseline MAP ranged from 79.67–104 mmHg, whereas peak MAP ranged from 93.33–126.67 mmHg.

**Table 2:**
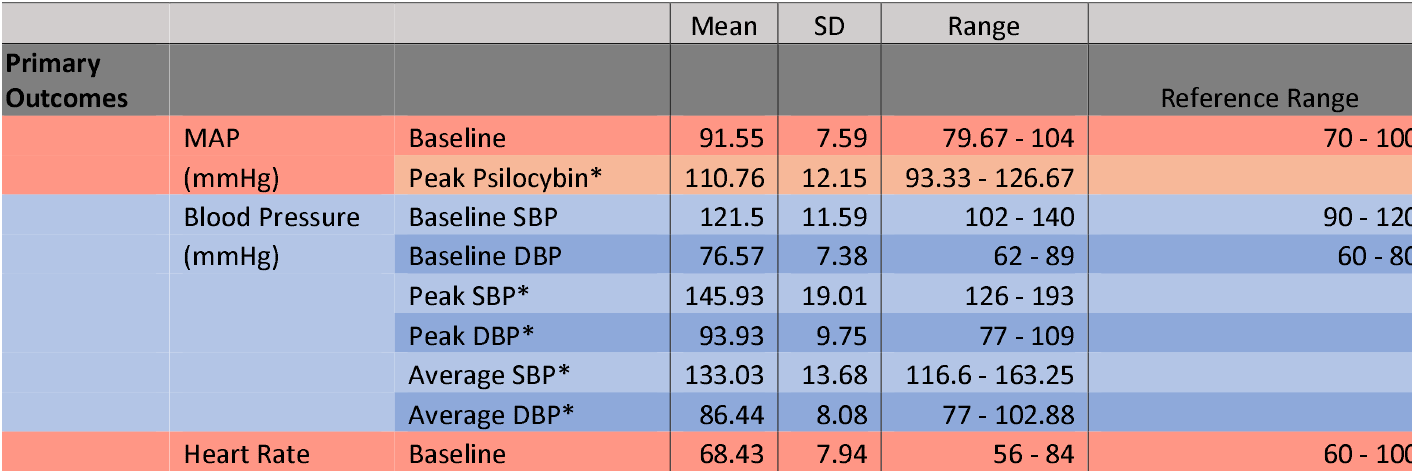
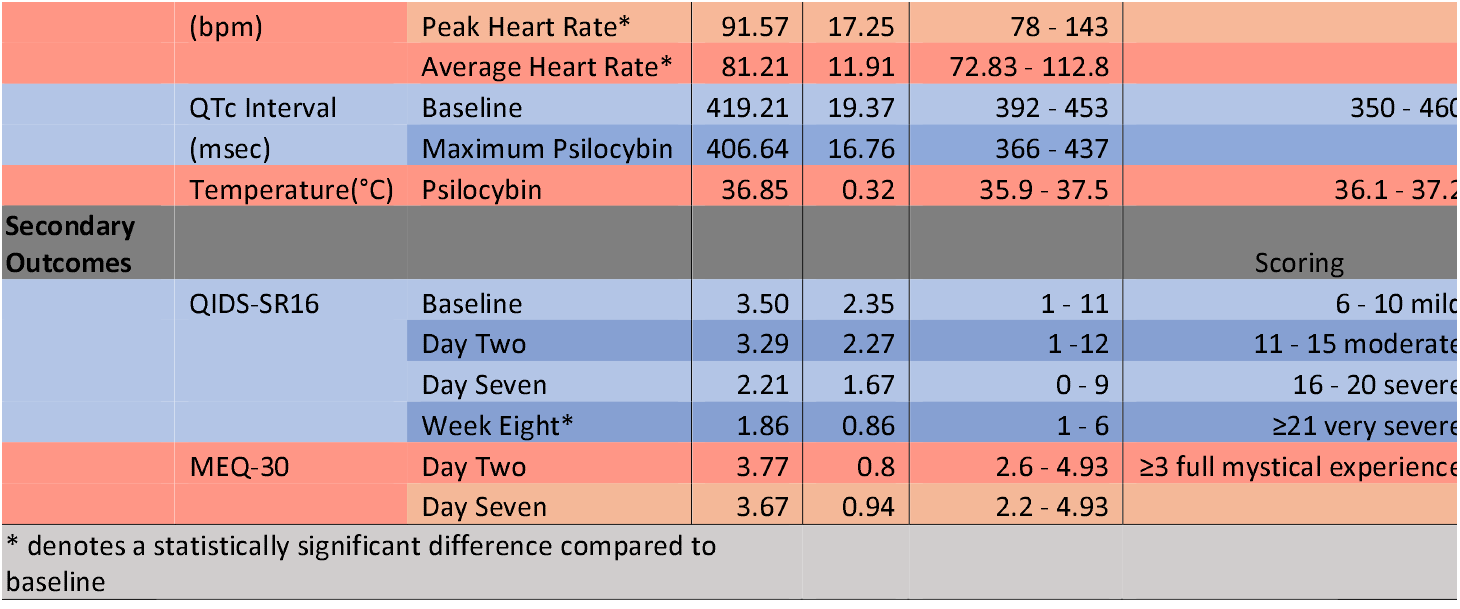
Summary of vital signs (primary outcome) and psychological questionnaires (secondary outcome).

**Figure 1:**
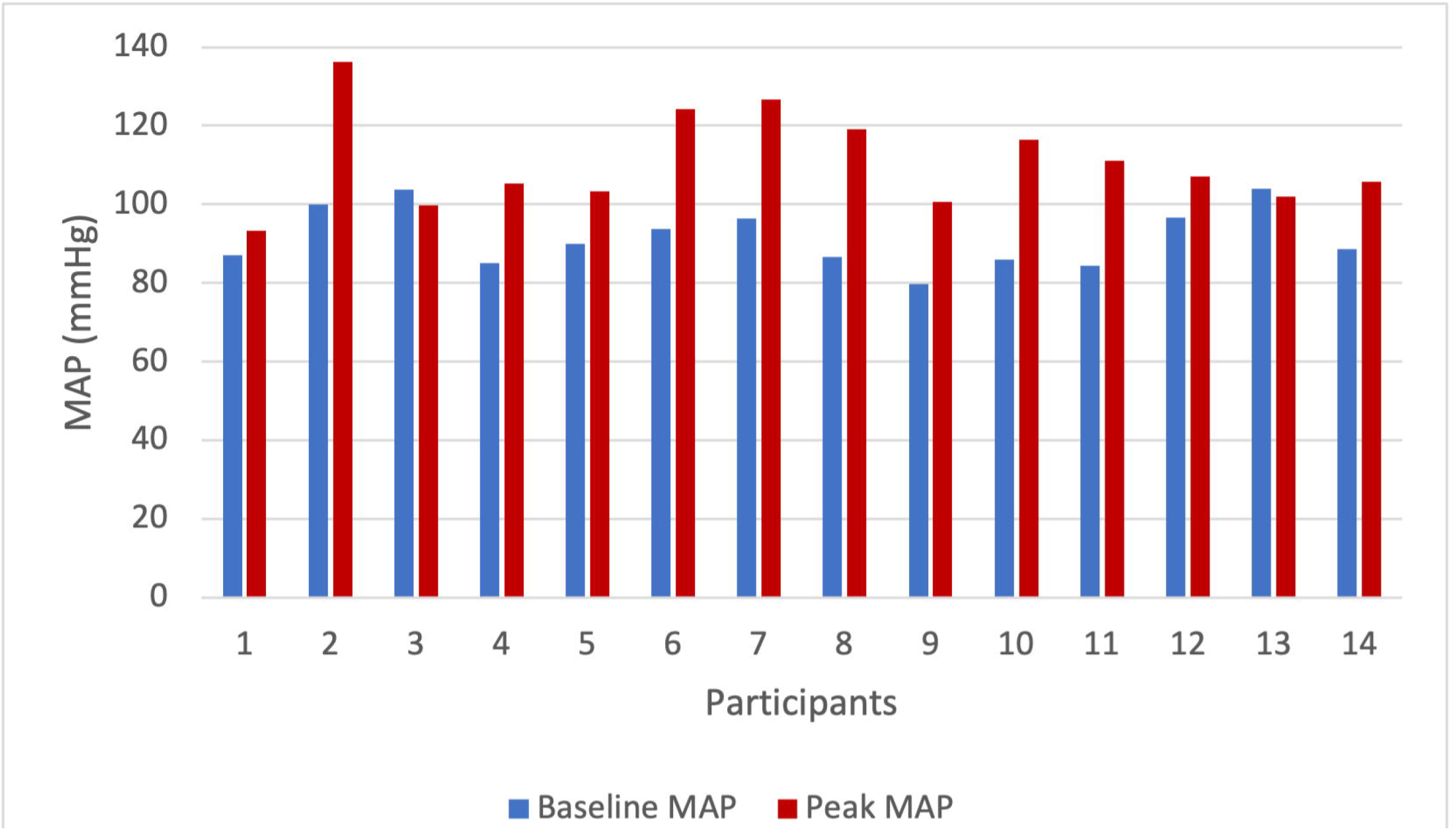
Comparison of baseline MAP to peak MAP during the psilocybin session for each participant.

A t-test for dependent means was conducted to assess if psilocybin had an effect on peak systolic blood pressure (SBP). The highest SBP for each participant during the psilocybin session was documented and these were compared to their baseline SBPs. Figure 2 compares baseline, average, and peak SBPs in each participant. There was a statistically significant effect *t*(13) = - 5.44, *p* < 0.001. Systolic blood pressure increased from an average baseline of 121.5 mmHg (*SD* = 11.59) to an average peak of 145.93 mmHg (*SD* = 19.01), demonstrating a likely effect of psilocybin on SBP. Baseline SBP ranged from 102-140 mmHg, while peak SBP ranged from 126-193 mmHg.

**Figure 2:**
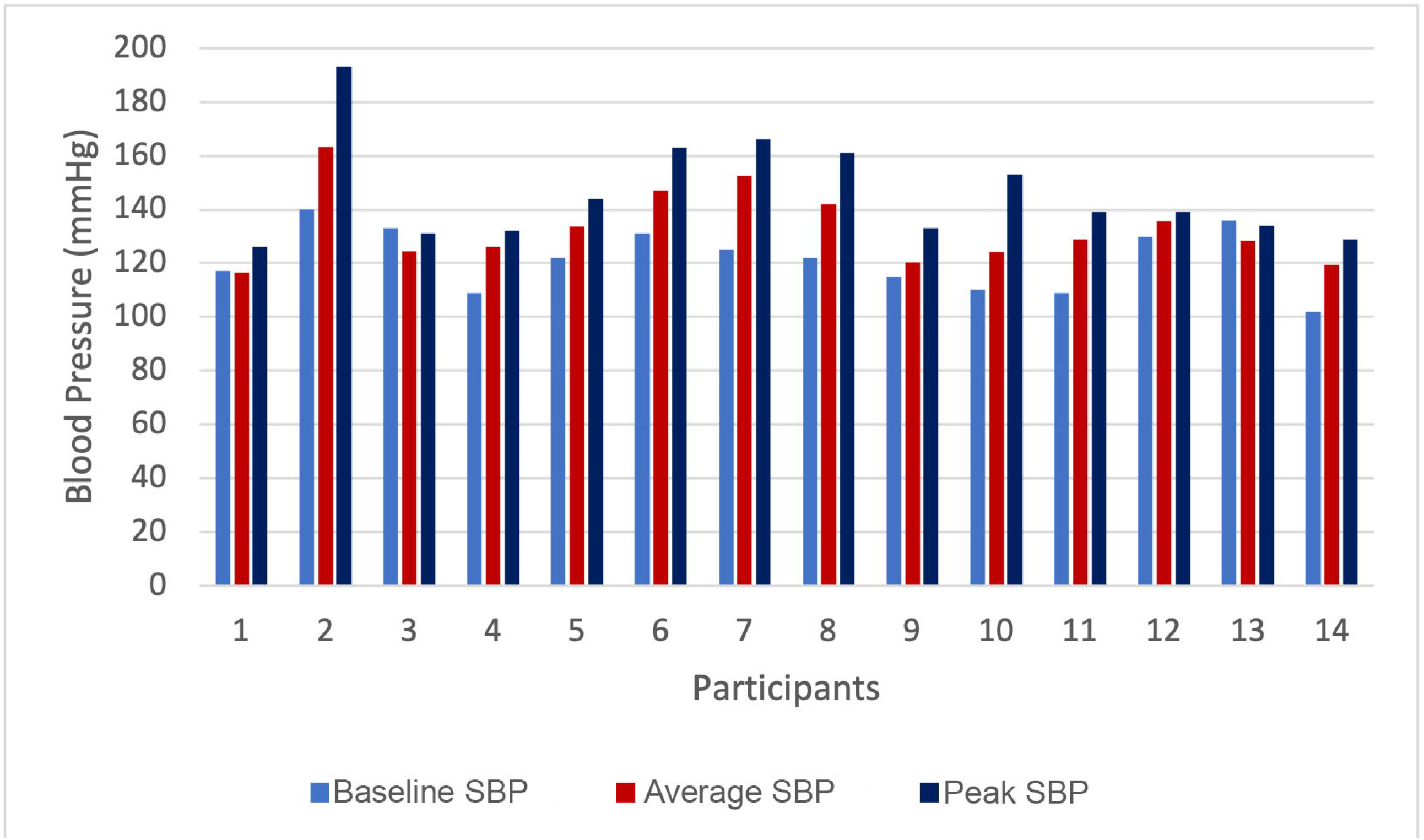
Comparison of each participant’s baseline SBP to the average and peak SBPs during the psilocybin session.

A t-test for dependent means was conducted to assess the effect that psilocybin had on peak diastolic blood pressure (DBP). The highest DBP for each participant during the psilocybin session was documented and these were compared to their baseline DBPs. Figure 3 compares baseline, average, and peak DBPs in each participant. There was a statistically significant effect *t*(13) = -5.54, *p* < 0.001. Diastolic blood pressure increased from an average baseline of 76.57 mmHg (*SD* = 7.38) to an average peak of 93.93 mmHg (*SD* = 9.75), demonstrating a likely effect of psilocybin on DBP. Baseline DBP ranged from 62-89 mmHg, while peak DBP ranged from 77-109 mmHg.

**Figure 3:**
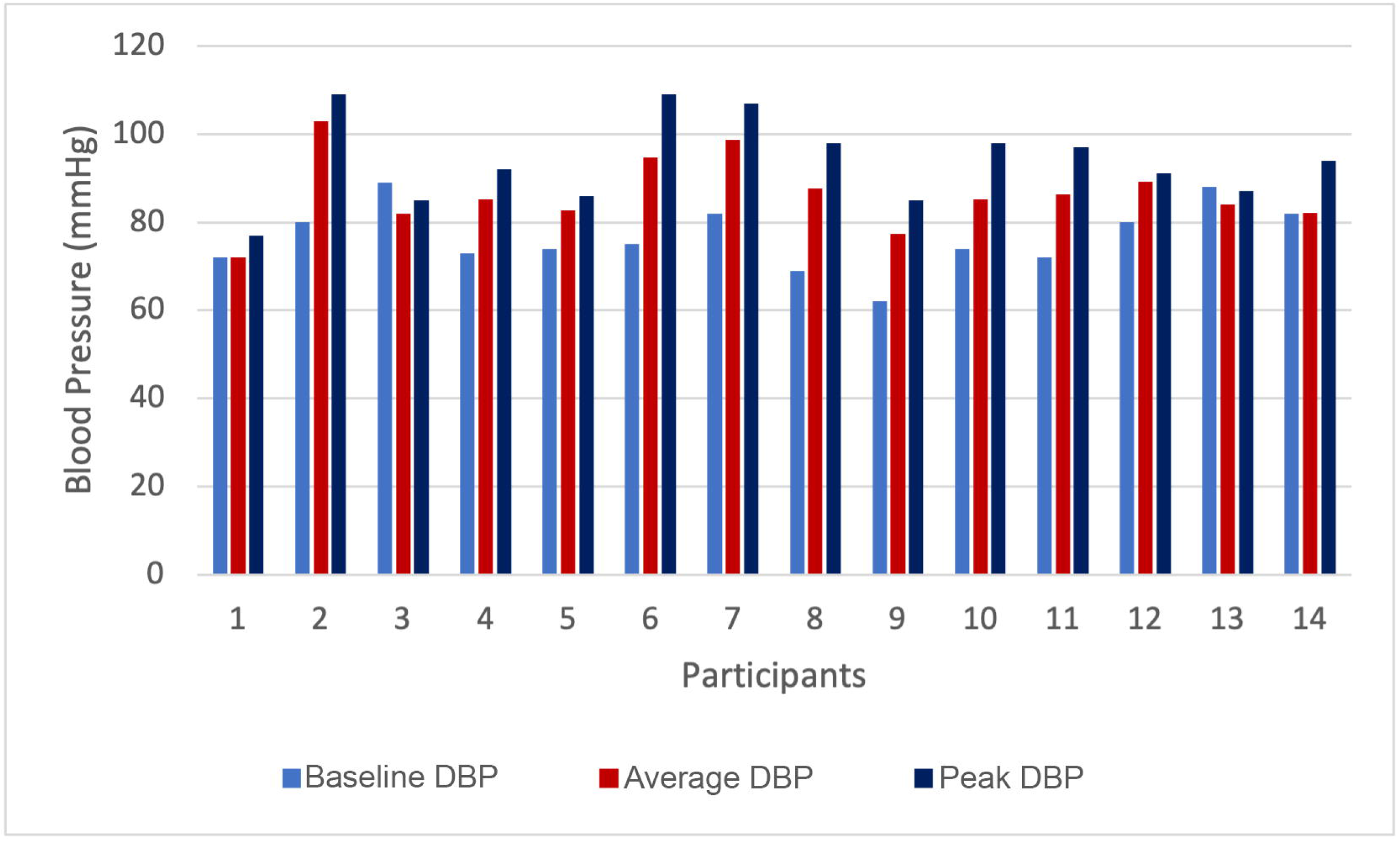
Comparison of each participant’s baseline DBP to the average and peak DBPs during the psilocybin session.

A t-test for dependent means was conducted to assess the effect that psilocybin had on average SBP. For each participant, all SBP data points collected during the psilocybin session were used to calculate an average SBP. These averages were compared to baseline SBP (Figure 2). There was a statistically significant effect *t(*13) = -3.88, *p* = 0.002. Systolic blood pressure increased from an average baseline of 121.5 mmHg (*SD* = 11.59) to an overall average of 133.03 mmHg (*SD* = 13.68); this is an average increase of 11.53 mmHg from baseline. Average SBP during the psilocybin session ranged from 116.6-163.25 mmHg.

A t-test for dependent means was conducted to assess the effect that psilocybin had on average DBP. For each participant, all DBP data points collected during the psilocybin session were used to calculate an average DBP. These averages were compared to baseline DBP (Figure 3). There was a statistically significant effect *t*(13) = -3.98, *p* = 0.002. Diastolic blood pressure increased from an average baseline of 76.57 mmHg (*SD* = 7.38) to an overall average of 86.44 mmHg (*SD* = 8.08); this is an average increase of 9.87 mmHg. Average DBP during the psilocybin session ranged from 77-102.88 mmHg.

A t-test for dependent means was conducted to examine the effect of psilocybin on peak heart rate. There was a statistically significant effect *t*(13) = -4.97, *p* < 0.001. Heart rate increased from a baseline of 68.43 bpm (*SD* = 7.94) to an average peak heart rate of 91.57 bpm (*SD* = 17.25), demonstrating a likely effect of psilocybin on heart rate. Baseline heart rate ranged from 56-84 bpm, while peak heart rate ranged from 78-143 bpm.

A t-test for dependent means was conducted to assess the effect of psilocybin on average heart rate. There was a statistically significant effect *t*(13) = -3.74, *p* = 0.002. Heart rate increased from an average baseline of 68.43 bpm (*SD* = 7.94) to an average of 81.21 bpm (*SD* = 11.91), demonstrating a likely effect of psilocybin on average heart rate. Average heart rate during the psilocybin session ranged from 72.83-112.8 bpm.

A t-test for dependent means was used to assess the effect of psilocybin on QTc interval. Psilocybin did not produce a significant effect on QTc interval *t*(13) = 2.15, *p* = 0.051, with an average baseline QTc interval of 419.21 (*SD* = 19.37) and an average maximum QTc interval of 406.64 (*SD* = 16.76). Baseline QTc intervals ranged from 392-453 and maximum QTc intervals during the psilocybin session ranged from 366-437.

Temperature ranged from 35.9-37.5°C during the psilocybin session, with a mean of 36.85 °C (*SD* = 0.32).

A one-way repeated measures ANOVA was used to assess the effect of psilocybin on depressive symptoms by assessing QIDS-SR16 scores before the psilocybin session and on day two, day seven, and week eight after the psilocybin session. There was a significant effect on QIDS-SR16 scores across the different time periods *F(*3, 39) = 3.80, *p* = 0.018. According to the Tukey HSD post hoc test, QIDS-SR16 scores after eight weeks (*M* = 1.86, *SD* = 0.86) were significantly lower than baseline scores 3.50 (*SD* = 2.35). No significant differences in scores were seen between baseline and day two, baseline and day seven, day two and day seven, day two and eight weeks, or day seven and eight weeks. Day two post-psilocybin QIDS-SR16 scores had a mean of 3.29 (S*D* = 2.27) and day seven post-psilocybin QIDS-SR16 scores had a mean of 2.21 (S*D* = 1.67).

A t-test for dependent means was conducted to determine if participants deemed their experience more mystical in nature two days after vs. seven days after the psilocybin session via the MEQ-30. There was not a significant difference seen between the two times *t(*13) = 1.35, *p* = 0.202. The mean MEQ-30 score for two days after the psilocybin session was 3.77 (SD = 0.80), and the mean MEQ-30 score for seven days after the psilocybin session was 3.67 (SD = 0.94). While there is not a significant difference between these times, both means are representative of a full mystical experience, which is determined by a score ≥ 3. On day two, the scores ranged from 2.6-4.93, and on day seven the scores ranged from 2.2-4.93. Overall, 11 of the 14 participants had a full mystical experience.

The adverse events experienced were generally mild, expected, and occurred within two days following the psilocybin session (Table 3). One participant experienced a severe headache during this time, but it was manageable with Tylenol and Advil. After seven days, the only adverse event experienced was mild anxiety by one participant, and no adverse events were present by the eight-week follow-up.

**Table 3:** Total number of adverse events (out of 14 participants) occurring two days, seven days, and eight weeks after the psilocybin session.

| Adverse Event | Mild | Moderate | Severe | Life-threatening | Fatal |
| --- | --- | --- | --- | --- | --- |
| 2 days post-psilocybin |  |  |  |  |  |
| Nausea | 3 |  |  |  |  |
| Vomiting |  |  |  |  |  |
| Headache | 2 | 3 | 1 |  |  |
| Anxiety | 1 |  |  |  |  |
| Confusion | 2 |  |  |  |  |
| Temporary fatigue or difficulty sleeping throughout the night | 3 | 1 |  |  |  |
| Anxiety, mania, or psychotic symptoms |  |  |  |  |  |
| Other |  |  |  |  |  |
| 7 days post-psilocybin |  |  |  |  |  |
| Nausea |  |  |  |  |  |
| Vomiting |  |  |  |  |  |
| Headache |  |  |  |  |  |
| Anxiety | 1 |  |  |  |  |
| Confusion |  |  |  |  |  |
| Fatigue |  |  |  |  |  |
| Anxiety, mania, or psychotic symptoms |  |  |  |  |  |
| Other |  |  |  |  |  |
| 8 weeks post-psilocybin |  |  |  |  |  |
| Nausea |  |  |  |  |  |
| Vomiting |  |  |  |  |  |
| Headache |  |  |  |  |  |
| Anxiety |  |  |  |  |  |
| Confusion |  |  |  |  |  |
| Fatigue |  |  |  |  |  |
| Anxiety, mania, or psychotic symptoms |  |  |  |  |  |
| Other |  |  |  |  |  |

## Discussion

The most significant finding from this safety analysis was the effect psilocybin has on blood pressure. Statistically significant elevations in both MAP, and systolic and diastolic blood pressure were seen. However, this observed increase in blood pressure and heart rate were expected adverse events, based on the known effects that classic psychedelics have on the cardiovascular system via certain serotonergic receptors (e.g., 5-HT_3_ and 5-HT_2A_) (Rossi, Hallak, Bouso Saiz, & Dos Santos, 2022). These receptors are not only directly related to the modulation of heart rate, but also affect vasoconstriction. While these events were expected, it was also predicted that such events would be transient and manageable in healthy participants, and this has been documented in other studies such as in Daniel & Haberman (2017) and Carbonaro, Johnson, Hurwitz, & Griffiths (2018).

While Hasler et al. (2004) did not find an overall significant main effect of psilocybin on MAP, they extended their analysis further to assess whether blood pressure increased at a particular time during psilocybin treatment; it was found that MAP was significantly elevated at 60 minutes post-administration. In a similar manner, DBP was only significantly elevated at 90 minutes post-administration. Yu et al. (2022) conducted a meta-analysis that analyzed psilocybin’s effect on the cardiovascular system as a secondary outcome. Compared with placebo, psilocybin treatment was associated with a significant increase in both systolic and diastolic blood pressure; an average increase of 19.00 mmHg and 8.66 mmHg was observed, respectively. This is comparable to the results of our study, in which there was an average increase in systolic and diastolic blood pressure from baseline by 11.53 mmHg and 9.87 mmHg, respectively. In a manner similar to the aforementioned studies, all 14 participants’ blood pressures and heart rates in our study returned to baseline levels as drug effects waned, and these levels remained normal at the eight-week follow-up appointments.

Another parameter of interest was the QTc interval. Dahmane, Hutson, & Gobburu (2021) conducted a study to determine the concentration-QTc relationship of psilocybin/psilocin. They determined that in the high dose group (ranging from 42-59 mg), at the time of C_max_, the upper limit of the 90% confidence interval of the mean ΔQTc exceeded the threshold level of regulatory concern (10 ms) at a psilocin concentration of 31.1 ng/mL. Such doses are much larger than the therapeutic psilocybin dose of 25 mg. At 25 mg, the mean psilocin C_max_ is about 18.7 ng/mL, with an associated ΔQTc of 2.1 ms (and 90% upper confidence level mean of 6.6 ms). Therefore, while psilocybin does have an effect on QTc interval, at therapeutic doses, the increase is not cause for clinical concern (Dahmane et al., 2021). Our study did not show a significant change in QTc interval, thus providing further evidence that, in healthy individuals, QTc prolongation is not a significant concern. The only atypical occurrence seen on three participants’ ECGs were ventricular extrasystoles, which resolved by the following reading. These are not considered clinically significant, as ventricular extrasystoles only become a cause for concern if they occur frequently or are symptomatic. No ventricular extrasystoles were seen on the eight-week follow-up ECGs.

Clearly, some individuals will experience blood pressure elevations during psilocybin use that extend into the hypertensive range; however, because such elevations are transient and manageable, we do not consider this clinically significant. The cardiovascular system is resilient enough to endure this brief increase, similarly to which both systolic blood pressure and heart rate increase during aerobic exercise (Cohen & Townsend, 2007). This research is also supported by emergency medicine studies that show no adverse outcomes in individuals presenting with hypertension to the Emergency Department in the ensuing two years (McAlister et al., 2021). As described by Alley & Copelin (2022), in general, it is normally not necessary to treat hypertensive urgencies, and it is often advised against due to the danger of unnecessary rapid correction that could lead to hypoperfusion. Virtually all episodes of hypertension with psilocybin use are without end-organ damage and do not require treatment.

If hypertension presents with symptoms of end-organ damage, such as headache, dizziness, shortness of breath, chest pain, vomiting, or vision changes, then further evaluation is required (Alley & Copelin, 2022). End-organ damage secondary to hypertension generally requires intravenous treatment, and therefore, transfer to the Emergency Department is required; it would be advisable to give an oral antihypertensive agent while waiting for transport. However, for most patients who experience hypertension due to drug effect, blood pressure will rapidly correct itself, especially with the administration of a benzodiazepine if warranted. As seen with one of the participants in our study, blood pressure increased to 193/108 mmHg during an emotionally intense period; this participant did not show symptoms of end-organ damage, and as expected, blood pressure corrected itself to 157/99 mmHg after 25 minutes, without pharmacological intervention. This participant’s peak blood pressure is an outlier in this study, as demonstrated by Figure 1 and Figure 2.

For context, despite significant clinical research, there are no examples of significant adverse physiological events, and despite extensive and longstanding recreational use, there is very scant evidence of associated physiological adversity. Adverse physiological effects that have occurred are generally a result of the concomitant use of other recreational drugs, including alcohol. We would hypothesize that use of psilocybin in the clinical setting would further diminish the risk of adverse physical outcomes. The only case of psilocybin intoxication resulting in cardiovascular dysfunction that we could find in the literature is described by Borowiak, Ciechanowski, & Waloszczyk (1998). This case described an 18-year-old man who was hospitalized following seizures and cardiopulmonary arrest after consumption of *Psilocybe semilanceata* mushrooms *–* the species of *Psilocybe* mushrooms containing the highest psilocybin concentration. This individual reported frequent psilocybin use, and investigations eventually revealed Wolff-Parkinson-White syndrome, which presumably contributed to arrhythmia (SVT) and myocardial infarction following use of the drug in this case. This individual had frequently consumed an unknown amount of a potent species of mushroom without prior evaluation from his physician – such uncontrolled conditions would not occur in a clinical setting.

Therefore, it would seem that psilocybin has a remarkably benign cardiovascular safety profile, and we expect that apparently heathy individuals can safely tolerate the therapeutic use of psilocybin without laboratory investigations or complete physical examinations. This will increase the accessibility of this treatment for many patients, without significantly compromising safety. Blood pressure assessment is a simple screening measure to conduct for all individuals seeking psilocybin-assisted therapy; however, we discourage measuring blood pressure prior to psilocybin ingestion on the day of treatment for several reasons. First, blood pressure is easily affected by anxiety; it can add another layer of stress if the message is that the patient will not be able to proceed with the treatment session if his/her blood pressure is too high. Second, in terms of maximizing efficacy, the process should not be over-medicalized.

Some basic screening measures are appropriate for individuals with known cardiovascular disease, poor exercise tolerance, or on QTc prolonging medications. In the case of those on QTc prolonging medications or those with known QTc concerns, an ECG and electrolyte panel should be performed prior to psilocybin administration. Any patient with uncontrolled hypertension or exercise-induced myocardial ischemia should have these concerns treated prior to psilocybin administration. Bloodwork (sodium, potassium, bicarbonate, urea, creatinine, calcium, and magnesium) should be conducted in those with abnormal or borderline ECGs, on diuretics, or with malnourishment. While thorough screening is necessary to attenuate the risk of adverse events from occurring in the first place, practitioners should have rescue medication (e.g., labetalol, nitroglycerin, and lorazepam and/or diazepam) on hand in the unlikely event a hypertensive emergency (or intense anxiety/agitation) occurs. Thus far, research and historical experience does not suggest that antipsychotic medications have a role in the acute setting.

The secondary outcome of our study was to assess the psychological effects of psilocybin; the specific psychological outcomes assessed were mood and mystical experience. The QIDS-SR16 was administered to participants on four occasions to evaluate mood: two days before the psilocybin session to obtain a baseline score, as well as two days, seven days, and eight weeks after the psilocybin session. Although the mean scores on days two, seven, and week eight were all lower than baseline (*M* = 3.50, *SD* = 2.35), only week eight showed a statistically significant lower score (*M* = 1.86, *SD* = 0.86). The QIDS-SR16 is a self-administered questionnaire that evaluates depressive symptomatology and correlates with the nine DSM-IV symptom criteria for depression; it is scored from 0-27, with higher scores indicating more severe depressive symptoms. Scores from 0-5 indicate no depression, 6-10 indicate mild depression, 11-15 moderate depression, 16-20 severe depression, and 21-27 very severe depression. Although a statistically significant decrease was seen from baseline to week eight, it does not represent a clinically significant difference when the overall scoring of the questionnaire is considered. This scale is designed to measure depression, whereas only one out of 14 participants’ baseline scores crossed the threshold for depression (with a score of 10). The mean scores of 3.50 at baseline and 1.86 at week-eight both signify no depression. As such, a limitation of our study was that this questionnaire was not the most sensitive tool to assess mood in healthy participants. A questionnaire that assesses mental well-being, such as the Brief Index of Self-actualization (BISA) revised, is likely a more appropriate tool to use to assess healthy participants; measurements of self-actualization are based on the idea that mental well-being should consider more than just the absence of disease, and should examine the extent to which personal potential is realized and achieved (Sumerlin & Bundrick, 1998). However, given the decrease in QIDS-SR16 scores in participants not suffering from clinical depression, we hypothesize that a commensurate or greater decrease would be seen in those suffering from depressive disorders.

Despite the fact that the QIDS-SR16 was not a sensitive tool for the assessment of mood in healthy participants, there are several clinically useful conclusions that can be drawn from the scores observed in our study. Although only experienced by a minority of participants (n = 3), it is important for practitioners to recognize the possibility that depressive symptoms may temporarily increase during the first week following a psilocybin treatment. Therefore, practitioners should follow-up with their clients during this time to ensure their well-being. It is widely accepted that this is a fertile time period for integration of the psychedelic experience, and it is hypothesized that the associated neuroplastic effects of psilocybin may help instantiate the therapeutic effects of therapy during this timeframe. With therapy, the brain can retain the new neural connections that the psychedelic state has cultivated. Essentially, in addition to the psilocybin-induced insights and biological changes that occur as a result of the acute drug effects, psilocybin also primes the brain for learning and healing in the weeks immediately following psilocybin use.

The MEQ-30 was the tool used to evaluate another subjective effect induced by psilocybin – the mystical experience. It is a 30-question, self-report questionnaire that assesses four different factors of the mystical experience: mysticism, positive mood, transcendence of space and time, and ineffability. Each question is scored on a scale of 0-5 and the final score is an average of all 30 questions. A score ≥ 3 represents a full mystical experience. In studies to date, the mystical experience appears to be positively associated with the therapeutic effects of psilocybin, and it is ultimately the combination of both physiological modifications in the brain and emotionally meaningful insights that drive the transformational changes that the patient experiences (Gukasyan & Nayak, 2021).

In our study, the MEQ-30 was completed on days two and seven after the psilocybin session. Participants’ perception of their mystical experience (or lack thereof) was not significantly different between the two measurements. Only three of 14 participants did not score ≥ 3. The mystical experience is often determined by “set” (factors related to the individual’s mindset as they enter the psychedelic session, including idiosyncratic personality dynamics, mood, past experiences, and expectations of the psilocybin experience), “setting” (the environment in which the psilocybin session occurs, including the cultural, physical, and social environment), dose, and individual pharmacokinetic characteristics (Hartogsohn, 2016). A 25 mg dose of psilocybin is equivalent to approximately 4-5 g of dried *Psilocybe cubensis* mushrooms (Haden, 2020). We speculate that one of our participants is a fast metabolizer of psilocybin, due to the fact that this participant only had a 30-minute experience, characterized by relaxation but not by the typical psychedelic effects. Of note, while this participant did not have a mystical experience, an increase in well-being was still described.

During the eight-week follow-up appointments, participants described the common experience of feeling “better,” “happier,” and “lighter” after the psilocybin session, despite not having perceived themselves as anxious prior to the session. Participants suggested that they had previously become accustomed to their prior levels of anxiety, as they did not have a reference point for comparison. After experiencing improvements in their mental well-being following psilocybin use, many participants felt optimistic that they would experience a greater sense of well-being in the future.

## Data Availability

All data produced are available online at https://doi.org/10.6084/m9.figshare.22329433.v1

## Author Contributions

JB, MB, RB, LG contributed to the conception and design of the study. JB organized the database. JB performed the statistical analysis. JB wrote the first draft of the manuscript. JB, RB, IM wrote sections of the manuscript. All authors contributed to manuscript revision, and read and approved the submitted version.

## Acknowledgments

We would like to thank The Newly Institute for the use of their facilities.

We would like to thank Natalie Bergstrom and Debbie White for their involvement in participant intake and data collection.

## Data Availability Statement

The original datasets presented in the study are available in a publicly accessible repository. This data can be found here: https://doi.org/10.6084/m9.figshare.22329433.v1

